# A psychrometric model to predict the biological decay of the SARS-CoV-2 virus in aerosols

**DOI:** 10.1101/2020.11.29.20240408

**Authors:** Clive B. Beggs, Eldad J. Avital

## Abstract

There is increasing evidence that the 2020 COVID-19 pandemic has been influenced by variations in air temperature and humidity. However, the impact that these environmental parameters have on survival of the SARS-CoV-2 virus has not been fully characterised. Therefore an analytical study was undertaken using published data to develop a psychrometric model to predict the biological decay rate of the virus in aerosols. This revealed that it is possible to predict with a high degree of accuracy (R^2^ = 0.718, p<0.001) the biological decay constant for SARS-CoV-2 using a regression model with enthalpy, vapour pressure and specific volume as predictors. Applying this to historical meteorological data from London, Paris and Milan over the pandemic period, produced results which indicate that the average half-life of the virus in aerosols was in the region 13-21 times longer in March 2020, when the outbreak was accelerating, than it was in August 2020 when epidemic in Europe was at its nadir. As such, this suggests that changes in virus survivability due the variations in the psychrometric qualities of the air might influence the transmission of COVID-19.

## Introduction

There is increasing evidence that the COVID-19 pandemic may have a seasonal component that is influenced by environmental factors [1-6], similar to that observed for the influenza A virus [7-10]. For example, in Hubei, China, it has been found that a 1°C increase in the air temperature was associated with a 36-57% decrease in the daily COVID-19 cases when the relative humidity (RH) was in the range 67.0-85.5%. Similarly, a 1% increase in RH led to a 11-22% decrease in the daily confirmed cases when the air temperature was in the range 5.0-8.2 °C [1]. In Bangladesh, higher air temperatures and higher RH levels have also been significantly associated with a reduction in the transmission of COVID-19 [11]. Indeed, it has found worldwide that the virus tends to spread more in regions that are cooler, with an average temperature of 5-11°C, and drier, with an absolute humidity (AH) of 4-7 g/m^3^, suggesting that the SARS-CoV-2 virus might exhibit seasonal behaviour [12]. Collectively, these findings suggest that COVID-19 might behave similarly to influenza A, also an enveloped RNA virus, which is known to survive in aerosols for much longer when AH and vapour pressure (VP) levels are low [10] - something that is possibly due to the ordering of the phospholipid envelope contributing to viral stability, leading to increased transmissibility at lower air temperatures [13].

The reasons for the seasonal variations in the behaviour of COVID-19 are unclear, but may be related to variations in air temperature [1, 4, 14-16], humidity [1, 4, 16] and ultraviolet (UV) radiation from sunlight [2, 17-19], as well as immunological [5, 17] and behavioural [20] changes. However, because of the many confounding factors that can affect COVID-19 transmission, it is difficult to identify the extent to which each of these environmental factors has influenced the course of the epidemic. For example, while it is known that both higher air temperatures [1, 16] and increased UV-B radiation in sunlight [17-19] are associated with reduced COVID-19 transmission, because in many parts of the world higher air temperatures are closely associated with increased UV-B radiation it becomes difficult to distinguish between the two. The situation is further complicated by the fact that many researchers misunderstand the concept of RH, which is a ratio of vapour pressures rather than an absolute value. RH is actually the ratio of the observed VP to saturated vapour pressure (SP) expressed as a percentage and as such is strongly affected by temperature because air can hold much more moisture at higher temperatures compared with low temperatures. Unfortunately, researchers do not always appreciate this fact, with the result that it is sometimes reported that during the winter months the air is more humid than in the summer because the RH values are higher, when in fact the air is actually much drier in winter due to SP being considerably lower. Furthermore, many researchers have performed statistical analysis on the RH, failing to appreciate that because it is a ratio involving SP, it is actually a function of air temperature (see equation 1 in the methods section below) and therefore not an independent variable. As such, there is a risk that questionable conclusions may have been reached. Consequently, although a clear association exists between COVID-19 prevalence and air temperature and humidity, the precise nature of that association and the reasons for it are much less clear.

Given this confusion, we designed the study presented here using published experimental and meteorological data (both in the public domain) with the aim of quantifying the extent to which changes in the psychrometric quality of the air (i.e. changes associated with temperature and humidity) influence the biological survival of the SARS-CoV-2 virus in aerosols. We did this because it is now recognised that ‘far-field’ transmission of COVID-19 can occur due the inhalation of small aerosolised respiratory droplets that can remain suspended in the air for considerable periods of time [21, 22], especially in poorly ventilated room spaces [23]. Therefore, there is a need to better understand how long the SARS-CoV-2 virus can remain viable in aerosols and the extent to which variations in air temperature and humidity influence biological longevity.

## Methods

A search of the relevant scientific literature (i.e. published literature, pre-prints and relevant websites) was undertaken to identify published data relating to the survival of the SARS-CoV-2 virus in aerosols under various environmental conditions. Only experiments conducted in the dark were included in the study, with those conducted in the presence of UV light excluded. From each study, the reported biological decay constant, *k*, together with the mean air temperature and RH used during experimentation were extracted and compiled into a dataset. With regard to this, the survival of the virus can be computed using the following first order decay equation.

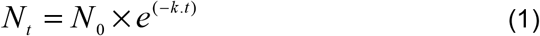

Where: *N*_*0*_ and *N*_*t*_ are the number of viable viral particles (virions) at time zero and *t* seconds respectively; and *t* is time in seconds.

From the reported mean air temperature and RH values, the psychrometric parameters saturated vapour pressure (SP), vapour pressure (VP), absolute humidity (AH), specific volume (SV), and specific enthalpy, *h*, were computed using the following empirical equations:

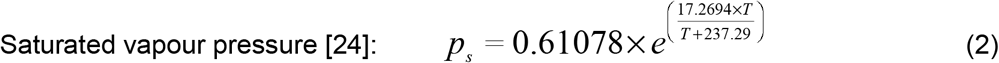

Where; *p*_*s*_ is saturated vapour pressure (kPa) and *T* is air temperature (°C).

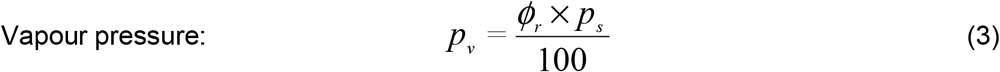

Where; *p*_*v*_ is saturated pressure (kPa) and *ϕ*_*r*_ is relative humidity (%).

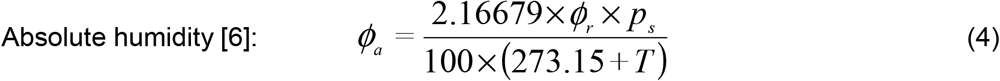

Where; *ϕ*_*a*_ is absolute humidity (kg of moisture per m^3^ of dry air).

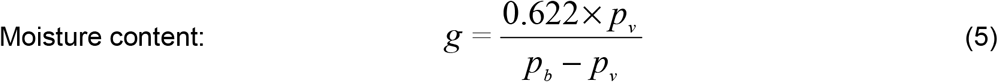

Where; *g* is moisture content (kg of moisture per kg of dry air) and *p*_*b*_ is barometric pressure (i.e. 101.325 kPa).

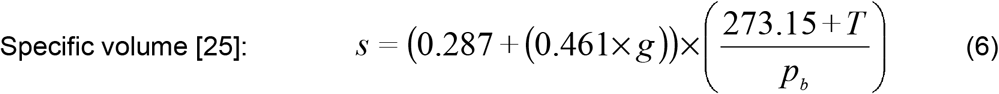

Where; *s* is the specific volume per kilogram of dry air (m^3^/kg)

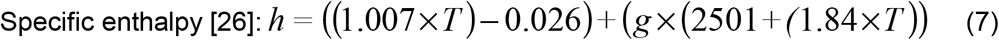

Where; *h* is specific enthalpy (kJ/kg).

Having computed the above psychrometric variables, partial correlation analysis was performed, controlling for whether or not the virus was aerosolised in artificial saliva. The impact of using artificial saliva on the *k* values was also assessed using a one-way ANOVA. All the statistical analysis was performed using R (R: A language and environment for statistical computing. R Foundation for Statistical Computing, Vienna, Austria), with p<0.05 deemed as significant.

Multiple linear regression analysis was then performed with the decay constant, *k*, as the response variable and SP, SV, enthalpy, and SV (or AH) as predictor variables. Because SV and AH are known to be highly correlated, it was necessary to build two competing models, one containing SV and the other, AH, in order to avoid multicollinearity issues. Similarly, RH was not included in the models because it is wholly described by VP and SP and therefore not independent. Refinement of the models was performed using backward exclusion, with only variables exhibiting p<0.1 retained. Heteroscedasticity was evaluated using the Breusch-Pagan test, and general applicability assessed using leave one out (LOO) cross-validation (CV).

In order to evaluate the impact of changes in psychrometric variables on the survival of the SARS-CoV-2 virus in aerosols the refined (minimum acceptable) regression model was used to predict weekly and monthly average *k* and biological half-life, *l*_*0*.*5*_, values from historical meteorological data [27] for London, Paris and Milan for the period 1^st^ January to 25^th^ October 2020. These were then compared with published COVID-19 case data acquired for the UK, France and Paris [28] for the period March to October, which approximates to the epidemic period in these countries. As it was only possible to acquire countrywide COVID-19 data, the aim here was simply to compare the relationship between the *k* values for the cities and the national case data, rather than to draw any firm inference. The half-life, *l*_*0*.*5*_, values used in this analysis were computed as follows.

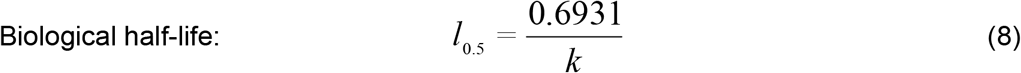

## Results

The results of the literature search are summarized in Table 1, which shows the biological decay constants, *k*, for the SARS-CoV-2 virus in aerosols, reported for 21 experiments by five research teams [29-33]. Of these, 13 experiments were conducted with the virus aerosolised in artificial saliva, while the rest used a buffer solution. No statistical difference was found (p = 0.546) between the *k* values reported for experiments conducted using artificial saliva and those that did not. Table 1 also includes computed psychrometric values for AH, VP, SP, enthalpy, and SV based on the mean air conditions reported for the various experiments.

**Table 1.** Reported biological decay constant, k, results for various air conditions, together with computed psychrometric values.

| Ref. No. | Artificial Saliva | Decay Constant, k, (min<sup>-1</sup>) | Mean Temp. (°C) | Mean Relative Humidity (%) | Absolute Humidity (kg/m<sup>3</sup>) | Vapour Pressure (kPa) | Saturated Vapour Pressure (kPa) | Specific Enthalpy (kJ/Kg) | Specific Volume (m<sup>3</sup>/kg) | Source |
| --- | --- | --- | --- | --- | --- | --- | --- | --- | --- | --- |
| 1 | No | 0.01060 | 22.0 | 65.0 | 0.0126 | 1.7185 | 2.6439 | 49.402 | 0.8504 | [29] |
| 2 | No | 0.00618 | 23.0 | 53.0 | 0.0109 | 1.4890 | 2.8094 | 46.728 | 0.8513 | [30] |
| 3 | Yes | 0.01000* | 20.0 | 20.0 | 0.0035 | 0.4676 | 2.3382 | 27.433 | 0.8342 | [31] |
| 4 | Yes | -0.00250* | 20.0 | 37.0 | 0.0064 | 0.8651 | 2.3382 | 33.708 | 0.8375 | [31] |
| 5 | Yes | 0.00800* | 20.0 | 53.0 | 0.0092 | 1.2393 | 2.3382 | 39.659 | 0.8406 | [31] |
| 6 | Yes | 0.01750* | 20.0 | 70.0 | 0.0121 | 1.6368 | 2.3382 | 46.031 | 0.8440 | [31] |
| 7 | No | 0.01500* | 20.0 | 20.0 | 0.0035 | 0.4676 | 2.3382 | 27.433 | 0.8342 | [31] |
| 8 | No | 0.01300* | 20.0 | 37.0 | 0.0064 | 0.8651 | 2.3382 | 33.708 | 0.8375 | [31] |
| 9 | No | 0.00750* | 20.0 | 53.0 | 0.0092 | 1.2393 | 2.3382 | 39.659 | 0.8406 | [31] |
| 10 | No | 0.01500* | 20.0 | 70.0 | 0.0121 | 1.6368 | 2.3382 | 46.031 | 0.8440 | [31] |
| 11 | Yes | -0.01100 | 10.0 | 20.0 | 0.0019 | 0.2456 | 1.2279 | 13.851 | 0.8040 | [32] |
| 12 | Yes | 0.01800 | 10.0 | 70.0 | 0.0066 | 0.8595 | 1.2279 | 23.451 | 0.8089 | [32] |
| 13 | Yes | 0.00600 | 20.0 | 20.0 | 0.0035 | 0.4676 | 2.3382 | 27.433 | 0.8342 | [32] |
| 14 | Yes | 0.01700 | 20.0 | 70.0 | 0.0121 | 1.6368 | 2.3382 | 46.031 | 0.8440 | [32] |
| 15 | Yes | -0.00300 | 30.0 | 20.0 | 0.0061 | 0.8486 | 4.2429 | 43.612 | 0.8659 | [32] |
| 16 | Yes | 0.06600 | 30.0 | 70.0 | 0.0212 | 2.9701 | 4.2429 | 78.197 | 0.8846 | [32] |
| 17 | Yes | 0.04000 | 40.0 | 20.0 | 0.0102 | 1.4751 | 7.3754 | 63.911 | 0.9001 | [32] |
| 18 | No | 0.00910 | 20.5 | 55.5 | 0.0099 | 1.3384 | 2.4116 | 41.755 | 0.8429 | [33] |
| 19 | Yes | 0.01590 | 20.5 | 55.0 | 0.0098 | 1.3264 | 2.4116 | 41.562 | 0.8428 | [33] |
| 20 | No | 0.02270 | 20.5 | 86.0 | 0.0153 | 2.0740 | 2.4116 | 53.614 | 0.8491 | [33] |
| 21 | Yes | 0.04000 | 20.5 | 81.0 | 0.0144 | 1.9534 | 2.4116 | 51.658 | 0.8481 | [33] |

\* Values estimated from plots.

The partial correlation results are presented in Table 2. These reveal that, after controlling for the use of artificial saliva, the *k* value was positively correlated with air temperature and RH, as well as with all the other psychrometric variables (all p<0.001). Notwithstanding this, the strongest correlations were with AH (r = 0.777), VP = 0.788; enthalpy (r = 0.794) and SV (r = 0.632). It can also be seen that the correlation between AH and VP was r = 0.999, implying that these two variables behave almost identically.

**Table 2.** Partial correlation r value results, controlling for artificial saliva.

|  | **k** | **Temp** | **RH** | **AH** | **Vap. Pres** | **Sat. pres** | **Enthalpy** | **Spec. vol** |
| --- | --- | --- | --- | --- | --- | --- | --- | --- |
| **k** | 1.000 | 0.491 | 0.474 | 0.777 | 0.788 | 0.478 | 0.794 | 0.632 |
| **Temp** | 0.491 | 1.000 | -0.173** | 0.392 | 0.423 | 0.965 | 0.753 | 0.971 |
| **RH** | 0.474 | -0.173** | 1.000 | 0.792 | 0.768 | -0.229* | 0.478 | 0.052** |
| **AH** | 0.777 | 0.392 | 0.792 | 1.000 | 0.999 | 0.312* | 0.900 | 0.601 |
| **Vap. Pres** | 0.788 | 0.423 | 0.768 | 0.999 | 1.000 | 0.346 | 0.915 | 0.628 |
| **Sat. pres** | 0.478 | 0.965 | -0.229* | 0.312* | 0.346 | 1.000 | 0.682 | 0.921 |
| **Enthalpy** | 0.794 | 0.753 | 0.478 | 0.900 | 0.915 | 0.682 | 1.000 | 0.889 |
| **Spec. vol** | 0.632 | 0.971 | 0.052** | 0.601 | 0.628 | 0.921 | 0.889 | 1.000 |

NB. All r values are significant at $p < 0.001$ , except: \* ( $p < 0.05$ ) and \*\* not significant.

The results of the linear regression analysis (utilising the predictor variables Temp, enthalpy, SP, SV, artificial saliva and VP or AH) are presented in Table 3, which shows two similar competing minimum adequate models, one containing VP and the other AH. These models produce almost identical results, although Model 1, utilizing VP, is the slightly superior model of the two, with: R^2^ = 0.718; mae = 0.007; and AIC = -129.9. The Breusch-Pagan test revealed no heteroscedasticity problems for either model. LOO CV analysis of Model 1 revealed the cross-validated R^2^ value to be 0.479 (mae = 0.009), implying that this model is general applicable. Interestingly, when the variables, enthalpy, VP, AH and SV were individually regressed onto the *k* value, the models produced were weaker, exhibiting for: enthalpy (R^2^ = 0.609); VP (R^2^ = 0.583); AH (R^2^ = 0.563); and SV (R^2^ = 0.397).

**Table 3.** Refined multiple linear regression models with the biological decay constant, *k*, as the response variable.

| **Model** | **Response Variable** | **Predictor Variables** | **Coefficient (b)** | **Significance (p value)** | **Model AIC (p value)** | **Model $R^2$ (mae)** | **LOO CV $R^2$ (mae)** |
| --- | --- | --- | --- | --- | --- | --- | --- |
| Model 1 | k | Intercept | 16.980 | 0.030 | -129.9 (<0.001) | 0.718 (0.007) | 0.479 (0.009) |
|  |  | Enthalpy | 0.062 | 0.028 |  |  |  |
|  |  | Vap. Pres. | -0.796 | 0.031 |  |  |  |
|  |  | Spec. Vol. | -21.950 | 0.030 |  |  |  |
| Model 2 | k | Intercept | 6.107 | 0.029 | -129.8 (<0.001) | 0.717 (0.007) | 0.451 (0.009) |
|  |  | Enthalpy | 0.022 | 0.025 |  |  |  |
|  |  | Abs. Humidity | -36.084 | 0.033 |  |  |  |
|  |  | Spec. Vol. | -7.882 | 0.029 |  |  |  |

The results for Model 1 are graphically presented in Figure 1, which shows a regression plot for the various studies aggregated together. From this, it can be seen that the *k* values from the various experiments all lie close (R^2^ = 0.718) to a linear plot line produced by Model 1.

**Figure 1.**
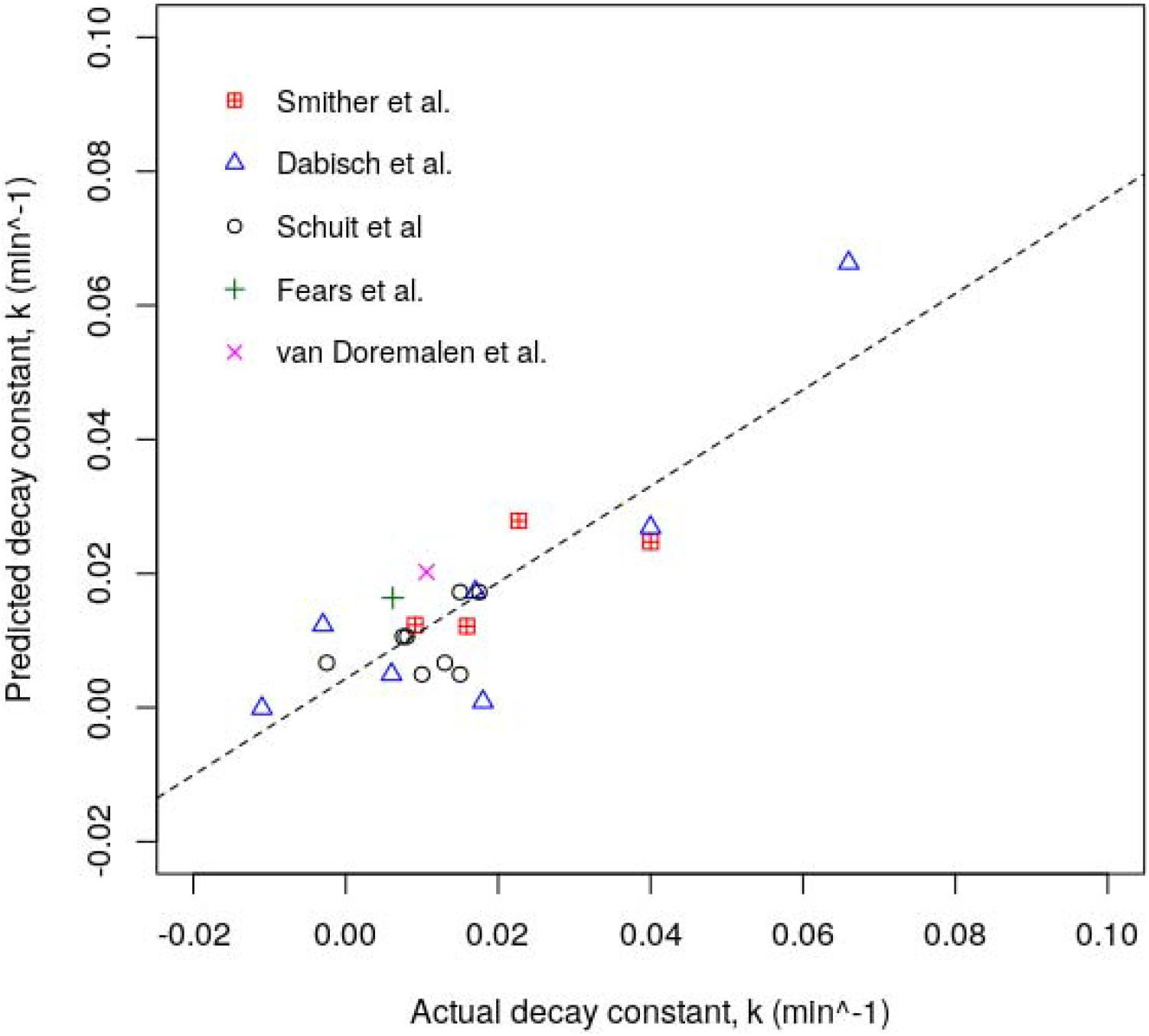
Scatter plot of actual and predicted k values for the various experiments.

The computed mean monthly psychrometric data for the period 1^st^ January to 25^th^ October 2020 for London, Paris and Milan are summarised in Table 4, whereas Table 5 shows the predicted mean monthly *k* and half-life values produced by applying Model 1 to the data in Table 4. From this it can be seen that the predicted biological half-life of the SARS-CoV-2 virus in aerosols during the winter months and early spring is much longer than that for the summer months. For example, in Milan during March, when the epidemic first took hold in Italy, the mean biological half-life of the virus was 517.2 minutes, whereas in August it was only 25.8 minutes – an approximate 20-fold reduction. A similar pattern was also observed for London and Paris.

**Table 4.** Average monthly values for the various psychrometric parameters for London, Paris and Milan during the 2020 (January to October).

| City | Parameter | January<br>Mean<br>(SD) | February<br>Mean<br>(SD) | March<br>Mean<br>(SD) | April<br>Mean<br>(SD) | May<br>Mean<br>(SD) | June<br>Mean<br>(SD) | July<br>Mean<br>(SD) | August<br>Mean<br>(SD) | September<br>Mean<br>(SD) | October*<br>Mean<br>(SD) |
| --- | --- | --- | --- | --- | --- | --- | --- | --- | --- | --- | --- |
| London | Temp (°C) | 7.78<br>(2.80) | 7.99<br>(2.89) | 8.00<br>(2.95) | 12.27<br>(4.52) | 15.09<br>(5.25) | 17.02<br>(4.37) | 18.23<br>(3.67) | 19.93<br>(4.80) | 16.20<br>(4.19) | 11.87<br>(2.47) |
| London | RH (%) | 83.25<br>(7.92) | 76.46<br>(10.84) | 70.26<br>(15.35) | 62.55<br>(17.13) | 56.26<br>(16.67) | 66.99<br>(15.98) | 66.05<br>(17.73) | 69.56<br>(16.83) | 71.83<br>(15.89) | 83.28<br>(10.60) |
| London | S.pres (kPa) | 1.0734<br>(0.2007) | 1.0905<br>(0.2111) | 1.0925<br>(0.2250) | 1.4854<br>(0.4703) | 1.8037<br>(0.6253) | 2.0089<br>(0.6129) | 2.1454<br>(0.5608) | 2.4247<br>(0.8051) | 1.9012<br>(0.5368) | 1.4067<br>(0.2311) |
| London | V.pres (kPa) | 0.8919<br>(0.1837) | 0.8320<br>(0.2000) | 0.7559<br>(0.1986) | 0.8664<br>(0.1536) | 0.9391<br>(0.2108) | 1.2776<br>(0.2478) | 1.3501<br>(0.2711) | 1.5923<br>(0.3296) | 1.3118<br>(0.2841) | 1.1608<br>(0.1922) |
| London | Sp.vol (m³/kg) | 0.8028<br>(0.0093) | 0.8029<br>(0.0096) | 0.8024<br>(0.0093) | 0.8154<br>(0.0134) | 0.8241<br>(0.0159) | 0.8324<br>(0.0136) | 0.8365<br>(0.0109) | 0.8434<br>(0.0153) | 0.8303<br>(0.0134) | 0.8167<br>(0.0082) |
| London | AH (kg/m³) | 0.0069<br>(0.0014) | 0.0064<br>(0.0015) | 0.0058<br>(0.0015) | 0.0066<br>(0.0011) | 0.0071<br>(0.0015) | 0.0095<br>(0.0018) | 0.0100<br>(0.0020) | 0.0118<br>(0.0024) | 0.0098<br>(0.0021) | 0.0088<br>(0.0014) |
| London | Enthalpy (kJ/kg) | 21.710<br>(5.552) | 20.991<br>(5.739) | 19.805<br>(5.277) | 25.869<br>(5.809) | 29.899<br>(7.445) | 37.239<br>(7.141) | 39.630<br>(5.872) | 45.273<br>(8.726) | 36.953<br>(7.664) | 30.120<br>(5.064) |
| Paris | Temp (°C) | 6.85<br>(3.59) | 8.93<br>(3.08) | 9.06<br>(3.54) | 15.24<br>(5.08) | 16.68<br>(5.09) | 18.89<br>(4.89) | 20.95<br>(4.30) | 22.43<br>(5.46) | 18.84<br>(5.06) | 12.59<br>(2.71) |
| Paris | RH (%) | 84.18<br>(9.84) | 76.99<br>(10.71) | 66.70<br>(19.07) | 54.74<br>(18.55) | 55.57<br>(16.93) | 62.65<br>(17.20) | 55.47<br>(16.82) | 60.34<br>(18.74) | 64.52<br>(17.96) | 79.79<br>(11.35) |
| Paris | S.pres (kPa) | 1.0178<br>(0.2433) | 1.1646<br>(0.2429) | 1.1824<br>(0.2952) | 1.8132<br>(0.5900) | 1.9872<br>(0.6479) | 2.2769<br>(0.7703) | 2.5588<br>(0.7405) | 2.8534<br>(1.0471) | 2.2751<br>(0.7587) | 1.4789<br>(0.2701) |
| Paris | V.pres (kPa) | 0.8517<br>(0.2146) | 0.8937<br>(0.2186) | 0.7809<br>(0.2666) | 0.9243<br>(0.2544) | 1.0422<br>(0.3076) | 1.3250<br>(0.2405) | 1.3280<br>(0.2782) | 1.5728<br>(0.3405) | 1.3669<br>(0.2839) | 1.1676<br>(0.2105) |
| Paris | Sp.vol (m³/kg) | 0.7998<br>(0.0117) | 0.8061<br>(0.0103) | 0.8056<br>(0.0115) | 0.8244<br>(0.0153) | 0.8295<br>(0.0159) | 0.8382<br>(0.0148) | 0.8441<br>(0.0126) | 0.8504<br>(0.0165) | 0.8384<br>(0.0156) | 0.8188<br>(0.0090) |
| Paris | AH (kg/m³) | 0.0066<br>(0.0016) | 0.0069<br>(0.0016) | 0.0060<br>(0.0020) | 0.0069<br>(0.0019) | 0.0078<br>(0.0022) | 0.0098<br>(0.0017) | 0.0098<br>(0.0021) | 0.0115<br>(0.0025) | 0.0101<br>(0.0020) | 0.0088<br>(0.0015) |
| Paris | Enthalpy (kJ/kg) | 20.139<br>(6.766) | 22.915<br>(6.223) | 21.276<br>(6.908) | 29.815<br>(7.457) | 33.161<br>(8.579) | 39.906<br>(7.230) | 42.058<br>(6.325) | 47.516<br>(8.569) | 40.536<br>(8.112) | 30.970<br>(5.570) |
| Milan | Temp (°C) | 3.05<br>(4.54) | 7.04<br>(5.31) | 8.52<br>(4.96) | 13.44<br>(5.88) | 18.13<br>(4.21) | 20.41<br>(4.87) | 23.92<br>(4.15) | 24.47<br>(4.51) | 19.52<br>(5.25) | 12.43<br>(3.87) |
| Milan | RH (%) | 83.51<br>(19.18) | 66.13<br>(26.63) | 68.84<br>(23.47) | 60.61<br>(24.40) | 66.81<br>(20.94) | 70.89<br>(19.83) | 65.55<br>(17.35) | 64.43<br>(18.78) | 72.32<br>(19.07) | 81.10<br>(17.53) |
| Milan | S.pres (kPa) | 0.7967<br>(0.2689) | 1.0640<br>(0.3887) | 1.1679<br>(0.4049) | 1.6397<br>(0.5979) | 2.1448<br>(0.5579) | 2.4953<br>(0.7695) | 3.0501<br>(0.7659) | 3.1672<br>(0.8628) | 2.3745<br>(0.7617) | 1.4835<br>(0.3756) |
| Milan | V.pres (kPa) | 0.6231<br>(0.1267) | 0.6431<br>(0.2524) | 0.7469<br>(0.2373) | 0.8913<br>(0.3237) | 1.3426<br>(0.2911) | 1.6358<br>(0.2654) | 1.8981<br>(0.3204) | 1.9108<br>(0.3297) | 1.6198<br>(0.3693) | 1.1614<br>(0.2627) |
| Milan | Sp.vol (m³/kg) | 0.7872<br>(0.0135) | 0.7987<br>(0.0158) | 0.8037<br>(0.0149) | 0.8190<br>(0.0174) | 0.8361<br>(0.0124) | 0.8452<br>(0.0146) | 0.8575<br>(0.0129) | 0.8592<br>(0.0135) | 0.8425<br>(0.0169) | 0.8183<br>(0.0122) |
| Milan | AH (kg/m³) | 0.0049<br>(0.0010) | 0.0050<br>(0.0019) | 0.0057<br>(0.0018) | 0.0067<br>(0.0024) | 0.0100<br>(0.0022) | 0.0121<br>(0.0019) | 0.0138<br>(0.0023) | 0.0139<br>(0.0024) | 0.0120<br>(0.0027) | 0.0088<br>(0.0019) |
| Milan | Enthalpy (kJ/kg) | 12.701<br>(5.862) | 17.062<br>(7.409) | 20.187<br>(7.157) | 27.474<br>(8.456) | 39.416<br>(6.463) | 46.451<br>(7.168) | 54.299<br>(7.479) | 55.071<br>(7.356) | 45.297<br>(9.823) | 30.708<br>(6.872) |

\* Data for 1 to 25<sup>th</sup> October 2020 only.

**Table 5.** Predicted monthly mean biological decay constant, k, and half-life values for London, Paris and Milan during the 2020.

| City | Parameter | January<br>Mean<br>(SD) | February<br>Mean<br>(SD) | March<br>Mean<br>(SD) | April<br>Mean<br>(SD) | May<br>Mean<br>(SD) | June<br>Mean<br>(SD) | July<br>Mean<br>(SD) | August<br>Mean<br>(SD) | September<br>Mean<br>(SD) | October*<br>Mean<br>(SD) |
| --- | --- | --- | --- | --- | --- | --- | --- | --- | --- | --- | --- |
| London | k.pred (min<sup>-1</sup>) | 0.00113<br>(0.00192) | 0.00108<br>(0.00183) | 0.00078<br>(0.00158) | 0.00262<br>(0.00265) | 0.00484<br>(0.00388) | 0.00963<br>(0.00528) | 0.01150<br>(0.00486) | 0.01697<br>(0.00828) | 0.00980<br>(0.00607) | 0.00488<br>(0.00355) |
| London | Mean half-life (min) | 613.4 | 641.8 | 888.6 | 264.5 | 143.2 | 72.0 | 60.3 | 40.8 | 70.7 | 142.0 |
| Paris | k.pred (min<sup>-1</sup>) | 0.00110<br>(0.00192) | 0.00171<br>(0.00254) | 0.00140<br>(0.00229) | 0.00500<br>(0.00359) | 0.00704<br>(0.00548) | 0.01143<br>(0.00562) | 0.01287<br>(0.00530) | 0.018.23<br>(0.00830) | 0.01224<br>(0.00639) | 0.00545<br>(0.00380) |
| Paris | Mean half-life (min) | 630.1 | 405.3 | 495.1 | 138.6 | 98.5 | 60.6 | 53.9 | 38.0 | 56.6 | 127.2 |
| Milan | k.pred (min<sup>-1</sup>) | 0.00008<br>(0.00035) | 0.00077<br>(0.00149) | 0.00134<br>(0.00208) | 0.00442<br>(0.00386) | 0.01142<br>(0.00521) | 0.01756<br>(0.00722) | 0.02626<br>(0.00870) | 0.02686<br>(0.00824) | 0.01762<br>(0.00800) | 0.00565<br>(0.00417) |
| Milan | Mean half-life (min) | 8663.8 | 900.1 | 517.2 | 156.8 | 60.7 | 39.5 | 26.4 | 25.8 | 39.3 | 122.7 |

\* Data for 1 to 25<sup>th</sup> October 2020 only.

When the mean weekly *k* values for London, Paris and Milan are compared with the COVID-19 case data for the UK, France and Italy (Figure 2), it can be seen that there is a broadly inverse relationship between the two plots (March to October, r = -0.258 (London); -0.124 (Paris); -0.512 (Milan)), with infections lowest during the summer months when the biological decay constant, *k*, is at its greatest. However, this relationship only reached significance in the case of Milan and Italy (p = 0.002).

**Figure 2.**
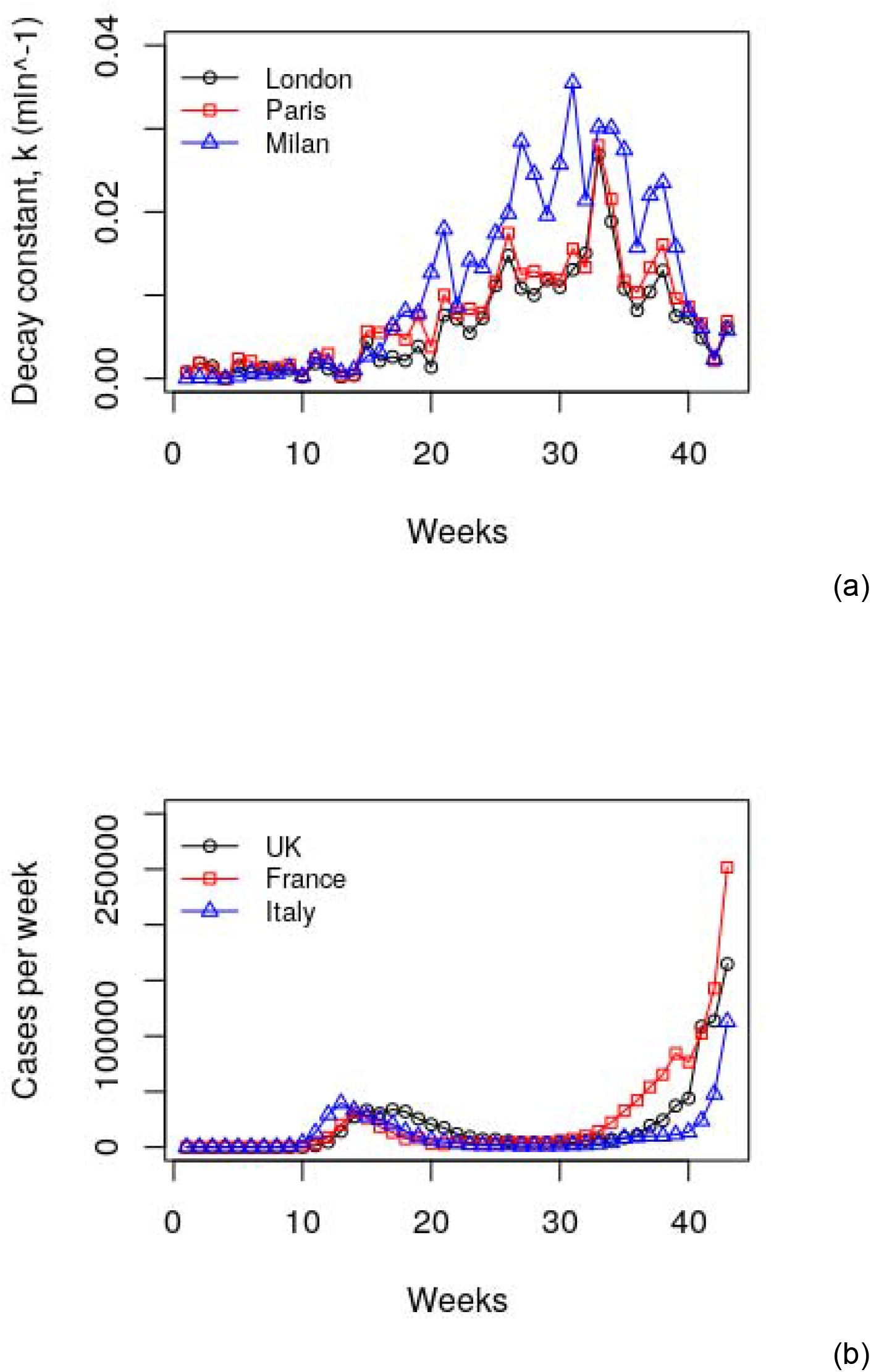
Plots of: (a) the biological decay constant, *k*, for London, Paris and Milan during the period 1^st^ January to 25^th^ October 2020.; (b) weekly COVID-19 cases for the UK, France and Italy during the same period.

## Discussion

The principal finding of our study is that the biological decay constant, *k*, for the SAR-CoV-2 virus in aerosols can be predicted with reasonable accuracy using a linear regression model with the variables: enthalpy, SV and VP (or AH) as predictors. As such, this further supports the growing body of evidence that COVID-19 transmission is influenced by changes in temperature and humidity [1, 4, 14-16] and suggests that in this respect the SARS-CoV-2 virus behaves similarly to influenza A [10, 11], which is also an enveloped RNA virus. Indeed, Shaman and Kohn [10] and Marr et al [34] (re-analysing Harper’s [35] original data) both produced similar results to ours, demonstrating that the survival of the influenza virus in aerosols is inversely correlated with VP (or AH) and air temperature. This mirrors our finding that the *k* value is most strongly correlated (r = 0.794, P<0.001) with specific enthalpy, *h*, which is a composite measure representing total heat energy in the air and as such is a function of both VP and air temperature. In practical terms, this implies that survival of the SARS-CoV-2 virus in aerosols is strongly influenced by the total energy in the air (i.e. the sensible heat energy of the dry air and the latent heat energy in the evaporated moisture combined). Consequently, as air temperature, VP and enthalpy increase, so the value of *k* also increases, with the result that the biological half-life, *l*_*0*.*5*_, decreases and the virus survival time becomes shorter.

The reasons why survival of the SARS-CoV-2 virus should reduce as the air temperature and VP increase are poorly understood. However, the fact that other enveloped viruses such as influenza, respiratory syncytial virus (RSV) and human coronavirus (HCoV), all exhibit seasonal cycles similar to COVID-19 [5], suggests that structural changes in the phospholipid envelope and surface proteins due to variations in temperature and humidity may be responsible [10, 34]. It may be that low-temperature conditions promote the ordering of lipids in the viral envelope and that this contributes to viral stability [13]. However, this does not explain how RH can affect viral stability in respiratory droplets when the actual virions are not exposed to the ambient air and therefore not directly interacting with the water vapour [34]. It therefore appears likely that evaporation from the droplet surface plays a key role in determining survivability. This is because evaporation affects the chemistry of droplets, which in turn might affect the stability of any viral particles contained within [34].

The evaporation process is dominated by the difference between the SP at the droplet surface and the ambient air VP [36]. It also depends on the temperature difference between the droplet surface and the ambient air; with flow convective effects influenced by the Reynolds, Schmidt and Prandtl numbers playing an important role [37]. As such, the droplet evaporation process exhibits a complex dependency on both temperature and humidity, which is difficult to model. However, Shaman & Kohn [10] developed a simplistic evaporation model which related time and the rate of decrease in the droplet’s radius, d*r*/d*t*, with the ratio of the air vapour pressure deficit to the ambient air temperature, (P_S_ - P_V_)/(273.15 + T), where T is in the range –20°C to 40°C. As such, they were able to show that the rate at which volumetric change occurs in aerosol droplets, and by inference changes in the solute (i.e. salts, proteins, etc.) concentration, are largely influenced by VP and air temperature. In particular, changes in pH within the aerosol droplet that are induced by evaporation may trigger conformational changes of the surface glycoproteins in enveloped viruses and subsequently compromise their infectivity [38].

As well as having a biological effect, evaporation profoundly influences droplet size and thus the aerodynamic behaviour of any respiratory droplets exhaled. With respect to this, respiratory droplets can be classified as being either large (>100-125 μm in diameter) or small (<100-125 μm in diameter). With large droplets the sedimentation process dominates over the evaporation process, with the result that they tend to travel only short distances from the source due to their ballistic behaviour [36, 39]. By comparison, small droplets are dominated by evaporation in their initial stage, rapidly reducing in size to become semi-solid aerosol particles containing proteins and salts [34] or fully dry particles called droplet nuclei, both of which can be transported long distances on convection currents and widely distributed in room spaces [22]. While the former are largely unaffected by air humidity, the latter are profoundly affected by VP if the flow convective effects are small [39]. So as well as affecting virus survival in aerosol droplets, air temperature and humidity can also affect the aerodynamic behaviour of respiratory aerosols and thus the transmission of viral diseases [10]. Indeed, Shaman & Kohn [10] were able to show that increased evaporation produced smaller aerosol particles (i.e. the formation of greater numbers of droplet nuclei) that stay airborne for longer. However, no strong correlation was found between the ratio of vapour pressure deficit to temperature and the actual transmission of influenza, suggesting that increased production of airborne droplet nuclei in low-VP conditions was not the principle means by which AH modulates influenza transmission [10]. Rather, they found a much stronger statistically significant relationship between VP and virus survival, suggesting that the modulation of viral survivability in aerosols is the primary means through which VP affects airborne influenza transmission [10].

In order to evaluate how changes in weather during the 2020 COVID-19 pandemic might have affected the SARS-CoV-2 biological decay constant, *k*, and thus survival half-life we applied our regression model (Model 1) to historical meteorological data for London, Paris and Milan (Table 4). This revealed (Table 5) that for all three locations the virus survived in the air for much longer during the winter, autumn and spring compared with the summer. For example, in Milan during March 2020 (when the Italian COVID-19 epidemic started to accelerate) the predicted mean half-life of the virus was 517 minutes, whereas in July and August (when the Italian epidemic reached its low point) the mean half-life was just 26 minutes. Mirroring Shaman & Kohn’s findings for influenza [10], this suggests that changes in VP (and by inference AH) and air temperature may have contributed to the seasonal fluctuations observed in the COVID-19 epidemic, particularly in temperate regions. Indeed, when the predicted weekly mean *k* values for these cities are compared with the weekly number of COVID-19 cases for the UK, France and Italy respectively, it can be observed that a broadly inverse relationship exists in all three countries, with COVID-19 cases lowest when the *k* values are highest. While, comparisons between individual cities and whole countries can only ever be considered illustrative, it nonetheless may be indicative of a relationship between virus survival in aerosols and transmission of the disease.

While in this study we have been able to characterise the strong relationship that exists between the psychrometric qualities of the air and survival of the SARS-CoV-2 virus in aerosols, it is important to note that we cannot say to what extent this contributes to the overall transmission of COVID-19. This is because the seasonal variations that result in large changes in air temperature and VP also coincide with changes in: UV-B irradiation levels; population behaviour; building occupancy levels; and ventilation rates, as well as changes in the human immune system. For example, in temperate regions during the summer months, populations spend more time outdoors, as well as ventilating buildings to a greater extent [23], both factors that tend to inhibit spread of the COVID-19. Likewise, viral degradation due to UV-B radiation in sunlight is greatly increased during the summer months [19, 31, 40], as are vitamin D levels due to exposure to sunlight [17, 19]. Furthermore, the effect of low air temperatures and VP levels during winter on the respiratory tract should not be ignored. Dehydration due to inhaling dry cold air can cause the mucosal layer in the respiratory tract to become more viscous, immobilising the cilia, and reducing the body’s ability to clear pathogens from the airways during wintertime [5]. Notwithstanding this, our finding that the SARS-CoV-2 virus is likely to survive in aerosols for much longer during winter and early spring compared with summer supports the observations of many other researchers [14-16, 29, 31]. Importantly however, it should be noted that the half-life survival times of several hours predicted by our model for the winter, spring and autumn months in Table 5, are much longer than the period virions are likely to remain airborne in a typical room space. As such, this raises two intriguing questions:

i. Does the psychrometric quality of the air alter the viral load in aerosol droplets that are inhaled and does this influence the progression of the epidemic? And;
ii. Does the psychrometric quality of the air alter the survival time and viral load in respiratory droplets that impact on surfaces and hands, etc. and does this in any way influence the transmission of COVID-19?

While it would appear reasonable to assume that increased viral load is associated with an increase in the biological half-life, the answers to both these question are at this stage unknown, and further investigation will be required in order to determine whether or not changes in air temperature and VP significantly affect the transmission of COVID-19 by either route.

Although we have characterised a strong relationship between survival of the SARS-CoV-2 virus in aerosols and air enthalpy, VP and SV, we are aware that our study has several noticeable limitations. Chief amongst these is our reliance on a limited dataset collected by disparate researchers, using a variety of experimental procedures. Furthermore, when calculating the psychrometric parameters we used the mean temperature and RH values reported in the respective publications, which meant that we could not compute any margins of error. Notwithstanding this, because of the ensemble approach taken and the strength of the linear relationship observed in Figure 1 and Table 3, which we cross-validated, it suggests that our findings and conclusions are valid. Having said this, we recommend that our results be treated, as being indicative only and that further experimental work should be undertaken to refine our findings.

## Conclusions

In conclusion, we have been able to demonstrate that survival of the SARS-CoV-2 virus in aerosols is inversely related to both air temperature and VP, with survival greatly increasing during the winter months when the air is cooler and drier. More specifically, we have been able to show that it is possible to predict with a high degree of accuracy (R^2^ = 0.718) the biological decay constant, *k*, of SARS-CoV-2 using a regression model with enthalpy, VP and SV as predictors. When applied to historical meteorological data for the 2020 COVID-19 pandemic for London, Paris and Milan, the results suggest that the average half-life of the virus in aerosols was in the region 13-21 times longer in March, when the outbreak was accelerating, than it was in August when it was at its nadir. As such, this suggest that changes in virus survivability due the variations in the psychrometric qualities of the air might play an important contributory role in influencing the transmission of COVID-19.

## Data Availability

All the relevant data is included in the manuscript.

## Notes

### Competing Interest Statement

The authors have declared no competing interest.

### Funding Statement

No external funding was received.

### Author Declarations

Leeds Beckett University

## References

1. Qi H, Xiao S, Shi R, Ward MP, Chen Y, Tu W, et al. COVID-19 transmission in Mainland China is associated with temperature and humidity: A time-series analysis. Science of the Total Environment. 2020:138778.

2. Merow C, Urban MC. Seasonality and uncertainty in global COVID-19 growth rates. Proceedings of the National Academy of Sciences. 2020;117(44):27456–64.

3. Audi A, AlIbrahim M, Kaddoura M, Hijazi G, Yassine HM, Zaraket H. Seasonality of Respiratory Viral Infections: Will COVID-19 Follow Suit? Frontiers in Public Health. 2020;8:576.

4. Ma Y, Zhao Y, Liu J, He X, Wang B, Fu S, et al. Effects of temperature variation and humidity on the death of COVID-19 in Wuhan, China. Science of The Total Environment. 2020:138226.

5. Moriyama M, Hugentobler WJ, Iwasaki A. Seasonality of respiratory viral infections. Annual review of virology. 2020;7.

6. Hoque MM, Saima U, Shoshi SS. Correlation of Climate Factors with the COVID-19 Pandemic in USA. Biomedical Statistics and Informatics. 2020;5(3):65.

7. Hemmes JH, Winkler K, Kool SM. Virus survival as a seasonal factor in influenza and poliomyelitis. Nature. 1960;188(4748):430–1.

8. Chan PKS, Mok HY, Lee TC, Chu IMT, Lam WY, Sung JJY. Seasonal influenza activity in Hong Kong and its association with meteorological variations. Journal of medical virology. 2009;81(10):1797–806.

9. Shaman J, Karspeck A. Forecasting seasonal outbreaks of influenza. Proceedings of the National Academy of Sciences. 2012;109(50):20425–30.

10. Shaman J, Kohn M. Absolute humidity modulates influenza survival, transmission, and seasonality. Proceedings of the National Academy of Sciences. 2009;106(9):3243–8.

11. Haque SE, Rahman M. Association between temperature, humidity, and COVID-19 outbreaks in Bangladesh. Environmental science & policy. 2020;114:253–5.

12. Sajadi MM, Habibzadeh P, Vintzileos A, Shokouhi S, Miralles-Wilhelm F, Amoroso A. Temperature and latitude analysis to predict potential spread and seasonality for COVID-19. Available at SSRN 3550308. 2020.

13. Polozov IV, Bezrukov L, Gawrisch K, Zimmerberg J. Progressive ordering with decreasing temperature of the phospholipids of influenza virus. Nature chemical biology. 2008;4(4):248–55.

14. Riddell S, Goldie S, Hill A, Eagles D, Drew TW. The effect of temperature on persistence of SARS-CoV-2 on common surfaces. Virology Journal. 2020;17(1):1–7.

15. Chin AWH, Chu JTS, Perera MRA, Hui KPY, Yen H-L, Chan MCW, et al. Stability of SARS-CoV-2 in different environmental conditions. The Lancet Microbe. 2020;1(1):e10.

16. Wang J, Tang K, Feng K, Lv W. High temperature and high humidity reduce the transmission of COVID-19. Available at SSRN 3551767. 2020.

17. Whittemore PB. COVID-19 fatalities, latitude, sunlight, and vitamin D. American journal of infection control. 2020;48(9):1042–4.

18. Sfica L, Bulai M, Amihaesei V-A, Ion C, Stefan M. Weather conditions (with focus on UV radiation) associated with COVID-19 outbreak and worldwide climate-based prediction for future prevention. Aerosol and Air Quality Research. 2020;20(9):1862–73.

19. Tang L, Liu M, Ren B, Wu Z, Yu X, Peng C, et al. Sunlight ultraviolet radiation dose is negatively correlated with the percent positive of SARS-CoV-2 and four other common human coronaviruses in the US. Science of The Total Environment. 2020;751:141816.

20. Kanzawa M, Spindler H, Anglemyer A, Rutherford GW. Will coronavirus disease 2019 become seasonal? The Journal of infectious diseases. 2020;222(5):719–21.

21. Beggs CB. Is there an airborne component to the transmission of COVID-19?: a quantitative analysis study. medRxiv. 2020.

22. Miller SL, Nazaroff WW, Jimenez JL, Boerstra A, Buonanno G, Dancer SJ, et al. Transmission of SARS-CoV-2 by inhalation of respiratory aerosol in the Skagit Valley Chorale superspreading event. medRxiv. 2020.

23. RAMP. The ventilation of buildings and other mitigating measures for COVID-19: a focus on winter 2020. arXiv preprint arXiv:200912781. 2020.

24. Alduchov OA, Eskridge RE. Improved Magnus form approximation of saturation vapor pressure. Journal of applied meteorology. 1996;35(4):601–9.

25. Chartered Institution of Building Services E. Reference Data: CIBSE Guide C: Routledge; 2001.

26. The enthalpy of moist air [Internet]. TheEngineeringToolbox.com. 2020 [cited 17th Novemeber 2020]. Available from: https://www.engineeringtoolbox.com/enthalpy-moist-air-d_683.html.

27. Weather for 243 countries of the world [Internet]. rp5.ru. 2020 [cited 4th November 2020]. Available from: https://rp5.ru/.

28. COVID-19 Coronavirus data [Internet]. European Centre for Disease Prevention and Control. 2020 [cited 25th October 2020]. Available from: https://data.europa.eu/euodp/en/data/dataset/covid-19-coronavirus-data.

29. van Doremalen N, Bushmaker T, Morris DH, Holbrook MG, Gamble A, Williamson BN, et al. Aerosol and surface stability of SARS-CoV-2 as compared with SARS-CoV-1. New England Journal of Medicine. 2020.

30. Fears AC, Klimstra WB, Duprex P, Hartman A, Weaver SC, Plante KC, et al. Comparative dynamic aerosol efficiencies of three emergent coronaviruses and the unusual persistence of SARS-CoV-2 in aerosol suspensions. medRxiv. 2020.

31. Schuit M, Ratnesar-Shumate S, Yolitz J, Williams G, Weaver W, Green B, et al. Airborne SARS-CoV-2 is Rapidly Inactivated by Simulated Sunlight. The Journal of Infectious Diseases. 2020;(doi: 10.1093/infdis/jiaa334).

32. Dabisch P, Schuit M, Herzog A, Beck K, Wood S, Krause M, et al. The Influence of Temperature, Humidity, and Simulated Sunlight on the Infectivity of SARS-CoV-2 in Aerosols. Aerosol Science and Technology. 2020;(just-accepted):1–15.

33. Smither SJ, Eastaugh LS, Findlay JS, Lever MS. Experimental Aerosol Survival of SARS-CoV-2 in Artificial Saliva and Tissue Culture Media at Medium and High Humidity. Emerging Microbes & Infections. 2020:1–9.

34. Marr LC, Tang JW, Van Mullekom J, Lakdawala SS. Mechanistic insights into the effect of humidity on airborne influenza virus survival, transmission and incidence. Journal of the Royal Society Interface. 2019;16(150):20180298.

35. Harper GJ. Airborne micro-organisms: survival tests with four viruses. Epidemiology & Infection. 1961;59(4):479–86.

36. Mittal R, Ni R, Seo J-H. The flow physics of COVID-19. Journal of fluid Mechanics. 2020;894.

37. Kincaid DC, Longley TS. A water droplet evaporation and temperature model. Transactions of the ASAE. 1989;32(2):457–0462.

38. Yang W, Marr LC. Mechanisms by which ambient humidity may affect viruses in aerosols. Applied and environmental microbiology. 2012;78(19):6781–8.

39. Seminara G, Carli B, Forni G, Fuzzi S, Mazzino A, Rinaldo A. Biological fluid dynamics of airborne COVID-19 infection. Rendiconti Lincei Scienze fisiche e naturali. 2020:1–33.

40. Ratnesar-Shumate S, Williams G, Green B, Krause M, Holland B, Wood S, et al. Simulated sunlight rapidly inactivates SARS-CoV-2 on surfaces. The Journal of Infectious Diseases. 2020.

